# Epidemiology of 869,220 varicose vein surgeries over 12 years in Brazil: trends, costs and mortality rate

**DOI:** 10.1101/2021.08.03.21261223

**Authors:** Marcela Juliano Silva, Andressa Cristina Sposato Louzada, Marcelo Fiorelli Alexandrino da Silva, Maria Fernanda Cassino Portugal, Marcelo Passos Teivelis, Nelson Wolosker

## Abstract

2.

**Objectives:** to assess the total numbers of surgical procedures performed between 2008 and 2019 for the treatment of varicose veins in the Public Health System, which exclusively insures more than 160 million Brazilians, the distribution of surgeries over time, as well as its costs and mortality rates.

**Study design:** retrospective cross-sectional population-based study

**Materials and Methods:** public and open data referring to all surgeries to treat chronic venous disease between 2008 and 2019 were extracted from the database of the Brazilian Public Health System.

**Results:** In total, 869,220 surgeries were performed to treat chronic venous disease in public hospitals and outpatient clinics in Brazil, with an average rate of 4.5 surgeries per 10,000 inhabitants per year. From 2015 on, we observed a slight downward trend in the total number of procedures. The total amount reimbursed by the government was US$ 232,775,518.11. A total of 49 deaths were registered following varicose vein surgery, corresponding to a mortality rate of 0.0056%.

**Conclusions:** a total of 869,220 surgeries performed to treat chronic venous disease over twelve years, with an overall rate of 4.52 procedures per 10,000 inhabitants per year. The mortality rate was very low, 0.0056%.

## 5. Introduction

Chronic venous disease (CVD) is very prevalent, progressive^1^ and with relevant morbidity^2,3^, leading to a high social, economic, and health burden^4–7^.

The prevalence of CVD may be even greater than 50% of the population^8–10^ and both the prevalence and the severity of CVD are expected to increase as the population ages^11,12^. Impairment of the patient’s health and quality of life is evident in the advanced stages of the CVD, especially when they present with venous ulcers^13^. However, patients report poorer health-related quality of life even with uncomplicated CVD^3^.

Furthermore, CVD represents a high social and economic burden. In the United States, CVD leads to 2 million workdays missed each year and to early retirement^4^, with a nationwide health care burden of more than $3 billion per year^5^. In Brazil, between 2005 and 2014, social security expenses with temporary disability due to CVD increased 3.5-fold, and more than 27 million workdays were lost^6^.

The standard surgical treatment, which includes ligation and stripping of saphenous veins as well as varicose veins’ phlebectomies, is cost-effective^14–17^. Even though this is a well-established and widely performed procedure to treat such a prevalent and morbid disease, to our knowledge, only a few countries have nationwide studies analyzing procedure rates and mortality, namely: Japan, the United States of America (USA) and Portugal, all of three developed countries. In Japan, they observed 41,246 surgeries for the treatment of CVD in 2014, with zero 30-day mortality^18^. In the USA, a total of 48,615 surgeries for CVD was performed between 2005 and 2017, with a 30-day mortality rate of 0.02%^19^. In Portugal, 153,382 patients underwent surgery to treat CVD in public hospitals between 2004 and 2016, with an in-hospital mortality rate of 0.015%^20^.

We believe that only population-level data generate real-world evidence, better supporting health care analysis and planning. To that aim, series larger and more diversified than those mentioned above may be necessary, including data from developing countries as well, which are lacking in the current literature.

Therefore, the objective of the present study was to retrospectively assess the total numbers of surgical procedures performed between 2008 and 2019 for the treatment of CVD in the Brazilian Public Health System (SUS), which exclusively insures more than 160 million Brazilians, the distribution of the surgeries over time, as well as its costs and mortality rates.

## 6. Materials and methods

Data were retrieved from the TabNet platform^21^, a public health information application developed by DATASUS, the Health Informatics Department of the Brazilian Ministry of Health. The TabNet system provides open data regarding procedures performed within the Brazilian Public Health System, input by hospitals and outpatient clinics adequately accredited. This accreditation and the information registration are a prerequisite for governmental reimbursement for the procedures performed.

Information on surgeries to treat CVD was selected from 2008 to 2019 at the TabNet platform of Brazil. Data included geographic region, number of procedures performed, mortality within the index admission in the hospital or in the outpatient facility, and the amounts reimbursed by the Public Health System.

Two types of procedure were assessed, according to coding as established by the management system for procedures and medications of the Brazilian Public Health System - SIGTAP (Sistema de Gerenciamento da Tabela de Procedimentos, Medicamentos e OPM): bilateral surgical treatment for CVD (04.06.02.056-6) and unilateral surgical treatment for CVD (04.06.02.057-4).

All data were collected from public access sites through computer programs of automated content access (web scraping). These automated navigation codes were programmed in Python language (v. 2.7.13, Beaverton – Oregon – USA) using the Windows 10 Single Language operational system. The data collection, platform field selection, and table adjustment steps were performed using the selenium-webdriver packages (v. 3.1.8, Selenium HQ, several collaborators worldwide) and pandas (v. 2.7.13, Lambda Foundry, Inc. e PyData Development Team, New York, USA). The Mozilla Firefox browser (v. 59.0.2, Mountain – California – USA) and geckodriver webdriver (v 0.18.0, Mozilla Corporation, Bournemouth, England) were used.

Following collection and treatment, data were organized and grouped in spreadsheets using the Microsoft Office Excel 2016® (v. 16.0.4456.1003, Redmond – Washington – USA) software.

Reimbursement amounts in Brazilian Real (R$, the official Brazilian currency) were converted into US dollars (US$) using the exchange rate on December 31st, 2013 (US$1 = R$2.34), the median date between the first and last cases evaluated.

Standardized procedure rates were calculated by dividing the number of procedures by the years of study and by the population exclusively insured by SUS, and then multiplying the quotient per 10,000.

### Statistical analysis

trends were evaluated using linear regression. For all tests, a p-value less than or equal to 0.05 was considered statistically significant.

This study was approved by the Ethics Committee of the institution where it was carried out (CAAE: 35826320.2.0000.0071). All data provided by DATASUS and TabNet is open and de-identified. For this reason, the Institutional Review Board exempted informed consent.

### Data availability statement

data associated with the paper are available in DATASUS TabNet platform^21^.

## 7. Results

In total, 869,220 surgeries were performed to treat CVD in Brazilian public hospitals and outpatient clinics between 2008 and 2019. The number and frequency of CVD surgery throughout the years are shown in Table 1. Bilateral procedures were the most frequently performed, representing 61.68% of the surgeries.

**Table 1:** Absolute and relative frequencies of unilateral and bilateral surgeries for varicose veins between 2008 and 2019

| Year | Unilateral<br>Procedures |  | Bilateral<br>Procedures |  | TOTAL |
| --- | --- | --- | --- | --- | --- |
|  | n | (%) | n | (%) |  |
| 2008 | 27,484 | 38.08 | 44,688 | 61.92 | 72,172 |
| 2009 | 25,836 | 36.05 | 45,838 | 63.95 | 71,674 |
| 2010 | 25,278 | 33.66 | 49,817 | 66.34 | 75,095 |
| 2011 | 24,625 | 32.85 | 50,332 | 67.15 | 74,957 |
| 2012 | 24,262 | 33.07 | 49,109 | 66.93 | 73,371 |
| 2013 | 25,800 | 34.17 | 49,698 | 65.83 | 75,498 |
| 2014 | 30,181 | 38.10 | 49,031 | 61.90 | 79,212 |
| 2015 | 26,540 | 38.62 | 42,174 | 61.38 | 68,714 |
| 2016 | 29,492 | 42.79 | 39,431 | 57.21 | 68,923 |
| 2017 | 30,494 | 45.43 | 36,635 | 54.57 | 67,129 |
| 2018 | 35,400 | 47.23 | 39,549 | 52.77 | 74,949 |
| 2019 | 27,693 | 41.01 | 39,833 | 58.99 | 67,526 |
| **Total** | **333,085** | **38.32** | **536,135** | **61.68** | **869,220** |

The distribution of procedures rates per 10,000 inhabitants exclusively dependent on the public health system per year is shown in Graph 1. The overall average rate was 4.5 surgeries for CVD treatment per 10,000 populations per year. The year 2014 showed the highest rates. From 2015 on, we observe a slight downward trend in the surgery rates (p=0.054).

The total amount reimbursed by the government per CVD surgeries between 2008 and 2019 was US$ 232,775,518.11. The overall average investment per surgery was US$ 267.80.

Total procedures, total deaths, and mortality rates among Brazilian geographic regions are presented in Table 2. A total of 49 deaths within the index admission in the hospital or outpatient facility was registered following CVD surgery, representing an overall mortality rate of 0.0056%.

**Table 2:**
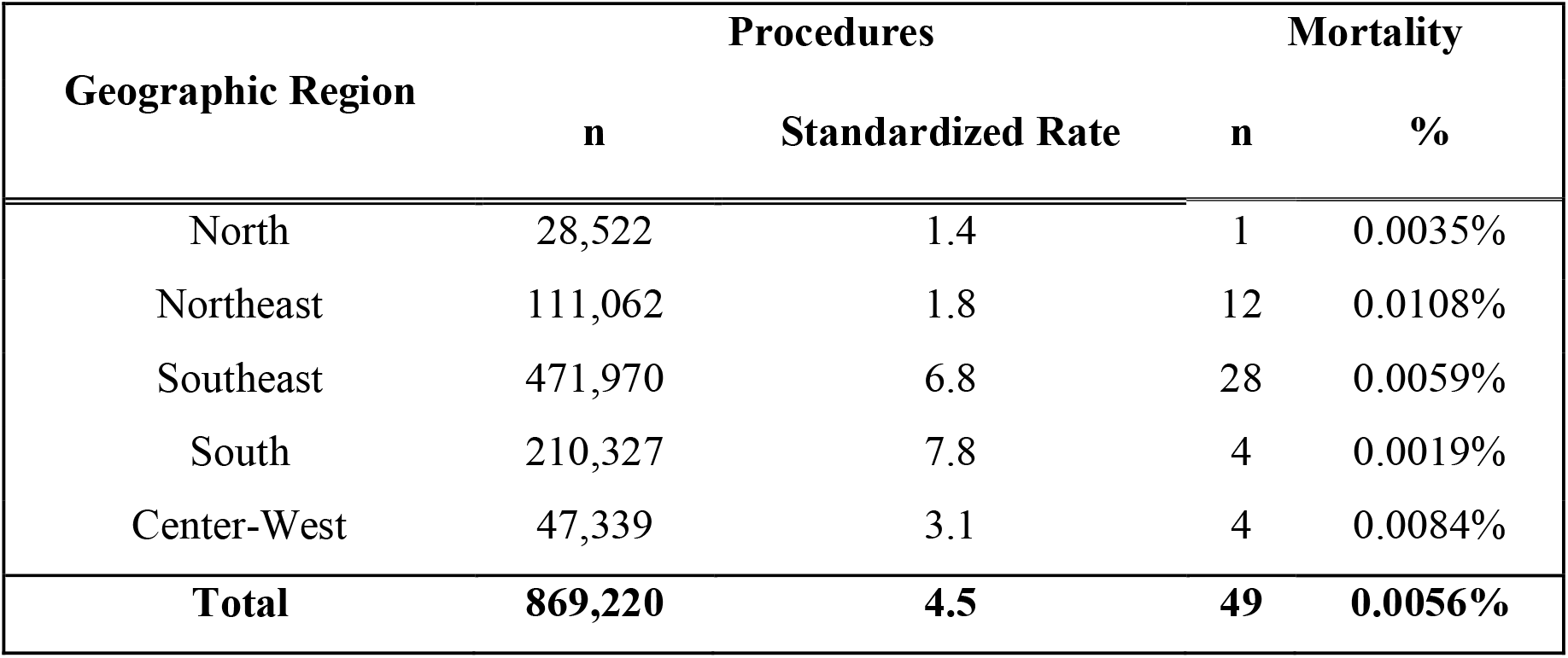
Absolute and relative mortality by geographic region for varicose vein surgery between 2008 and 2019

| Geographic Region | Procedures |  | Mortality |  |
| --- | --- | --- | --- | --- |
|  | n | Standardized Rate | n | % |
| North | 28,522 | 1.4 | 1 | 0.0035% |
| Northeast | 111,062 | 1.8 | 12 | 0.0108% |
| Southeast | 471,970 | 6.8 | 28 | 0.0059% |
| South | 210,327 | 7.8 | 4 | 0.0019% |
| Center-West | 47,339 | 3.1 | 4 | 0.0084% |
| **Total** | **869,220** | **4.5** | **49** | **0.0056%** |

The southeast region of Brazil performed more than half of the procedures and showed an intermediate mortality rate. Both the South and the Southeast had the higher standardized rates of CVD surgery per year per 10,000 populations, with rates more than 3 times higher than the ones observed in the North and Northeast.

## 8. Discussion

We observed a total of 869,220 surgeries to treat CVD, which corresponds to the largest series of the mostly performed vascular surgery.

Bilateral procedures were more frequent, which is likely a reflection of public reimbursement and logistics in Brazil, as waiting lines are frequently long.

The overall standardized rate was 4.5 surgeries for CVD treatment per 10,000 populations per year, which is similar to the ones observed in Japan^18^ and in the United States when considering the Medicare enrollees^22^, but at least 5 times smaller than the ones reported in some European countries such as Finland^23^, Belgian^24^, and Scotland^25^. Since the Brazilian prevalence of CVD is estimated to be high, 37.9% in men and 50.9% in women^26^, the 5-fold higher rates of surgical procedures in these European countries can not be explained by a higher prevalence in a similar fashion. Health care is mostly publicly funded in Brazil, Finland^27^, Belgium^28^ and Scotland^29^, therefore payment source can not explain the difference either. A possibility to be studied is that the population of these European countries is much smaller than that of Brazil, the United States of America and Japan, so perhaps the logistics of treating a low-mortality disease such as CVD are easier.

We also observed a disparity internally, as the South and Southeast, the two regions with the highest population density and development indexes, had CVD surgery rates more than 3 times higher than the ones observed in the North and Northeast, the poorest and least developed Brazilian regions^30^, which may suggest an impact of socioeconomic factors on the surgical treatment of CVD.

Regarding the distribution of surgeries over time, in Brazil, we observed a slight decrease in the total surgical procedures performed under the Public Health System since 2015. This finding contrasts with recent reports in the literature of a rising tendency to treat CVD surgically^18,24,25^. One possible explanation may be an increasing rate of ultrasound-guided sclerotherapy with polidocanol foam to treat CVD, an outpatient procedure that is also reimbursed by the Brazilian government but was not evaluated in our study. Concerning endovenous procedures, although they may be accounted for an upward trend in procedures performed to treat CVD in other countries currently^18,22^, they are unlikely to influence the standard surgery rates in Brazil, as SUS does not ordinarily reimburse them. Some patients may be seeking treatment for CVD in the private sector, as the waiting lists are often long in SUS, but these patients probably do not represent a large sample, since most Brazilians exclusively insured by the SUS cannot afford to pay surgical treatments out of their pockets.

The average spent by the government per procedure was US$ 267.80. The average amount paid by the governments of China^31^ and the United Kingdom^14^ per surgery is at least twice as much the amount paid by the Brazilian government. Due to the limitations of the data collection, we were not able to distinguish the amount paid for unilateral and bilateral surgeries, or for inpatient or outpatient procedures, we only have access to an average amount. Moreover, the government’s refund is based on a fixed compensation table and may not reflect the actual hospital and outpatient expenses, which is another limitation of our study.

After the 869,220 surgeries performed to treat CVD, we observed 49 deaths within the index admission in the hospital or in the outpatient facility, corresponding to a mortality rate of 0.0056%. To our knowledge, the only other population-based study for comparison was the one evaluating the data from Portuguese public hospitals, with an in-hospital mortality rate of 0.015%^20^. The mortality rate for CVD surgeries in Portugal was three-fold higher. A possible explanation for this difference is that our series is almost six-fold larger, maybe better reflecting real-life statistics. Another explanation is that in Portugal, the surgery rate for CVD is more than double the one in Brazil, thus it is possible that they encompass more at-risk patients in Portugal. Nonetheless, both mortality rates are very low, which underlines the safety of the surgical treatment.

Still regarding mortality assessment, some limitations of our study are noteworthy. As our data are anonymous, we do not know the causes of death and follow-up is not possible, therefore, deaths after discharge were missed, and we are unable to calculate 30-day mortality, as authors evaluated in Japan^18^ and USA^19^. Consequently, we may be observing more deaths related to anesthetics and bleeding complications and perhaps missing some pulmonary embolism-related deaths.

## Limitations

In addition to the limitations already mentioned: the inability to distinguish the amount paid for unilateral and bilateral surgeries and the surgeries which involved or not saphenous vein treatment, refund based on a fixed compensation table and the impossibility to follow up, some other limitations are worthy of note.

We only have data on total number of procedures performed, which possibly does not reflect the number of patients treated, as some patients may have undergone more than one procedure to treat CVD, such as two unilateral procedures or even re-interventions considering that the study period is large.

As inherent to a retrospective analysis using web scraping data collection, our study is limited by the loss of patient information and eventual miscoding of the administrative database, which should not be statistically significant, given the large size of our population.

Despite these limitations, this is a comprehensive analysis of the surgical treatment of CVD under the Brazilian Public Health System, with the largest series of the most frequently performed vascular surgery in the literature, granting real-life statistics with high external validity. Therefore we believe this study presents a valuable tool for a better understanding of the surgical management of CVD and highlights its safety in the real world.

## Conclusions

We observed a total of 869,220 surgeries performed to treat CVD over twelve years, with an overall rate of 4,52 procedures per 10,000 inhabitants per year. The mortality rate was very low, 0.0056%.

## Data Availability

All data are publicly available at DATASUS platform

## 4. Abbreviation List

CVD: Chronic venous disease
USA: United States of America
SUS: Brazilian Public Health System

## 9. Acknowledgements and Declarations

We thank Sergio Kuzniec, M.D. Ph.D. for his critical reading of the manuscript.

This study was approved by the Ethics Committee of the institution where it was carried out (CAAE: 35826320.2.0000.0071).

No author has conflict of interest, and this research did not receive any specific grant from funding agencies in the public, commercial or not-for-profit sectors.

All authors have seen and approved this manuscript.

## Authors’ contributions

Conception and design: MPT, NW

Data collection: MJS, ACSL, MFAS, MFCP

Analysis and interpretation: MJS, ACSL, MFAS, MFCP, MPT, NW

Writing the article: MJS, ACSL, MFAS, MFCP

Critical revision of the article: MJS, ACSL, MFAS, MFCP, MPT, NW

Final approval of the article: MJS, ACSL, MFAS, MFCP, MPT, NW

Overall responsibility: MJS, ACSL, MFAS, MFCP, MPT, NW

## 12. Figure legends

**Graph 1.**
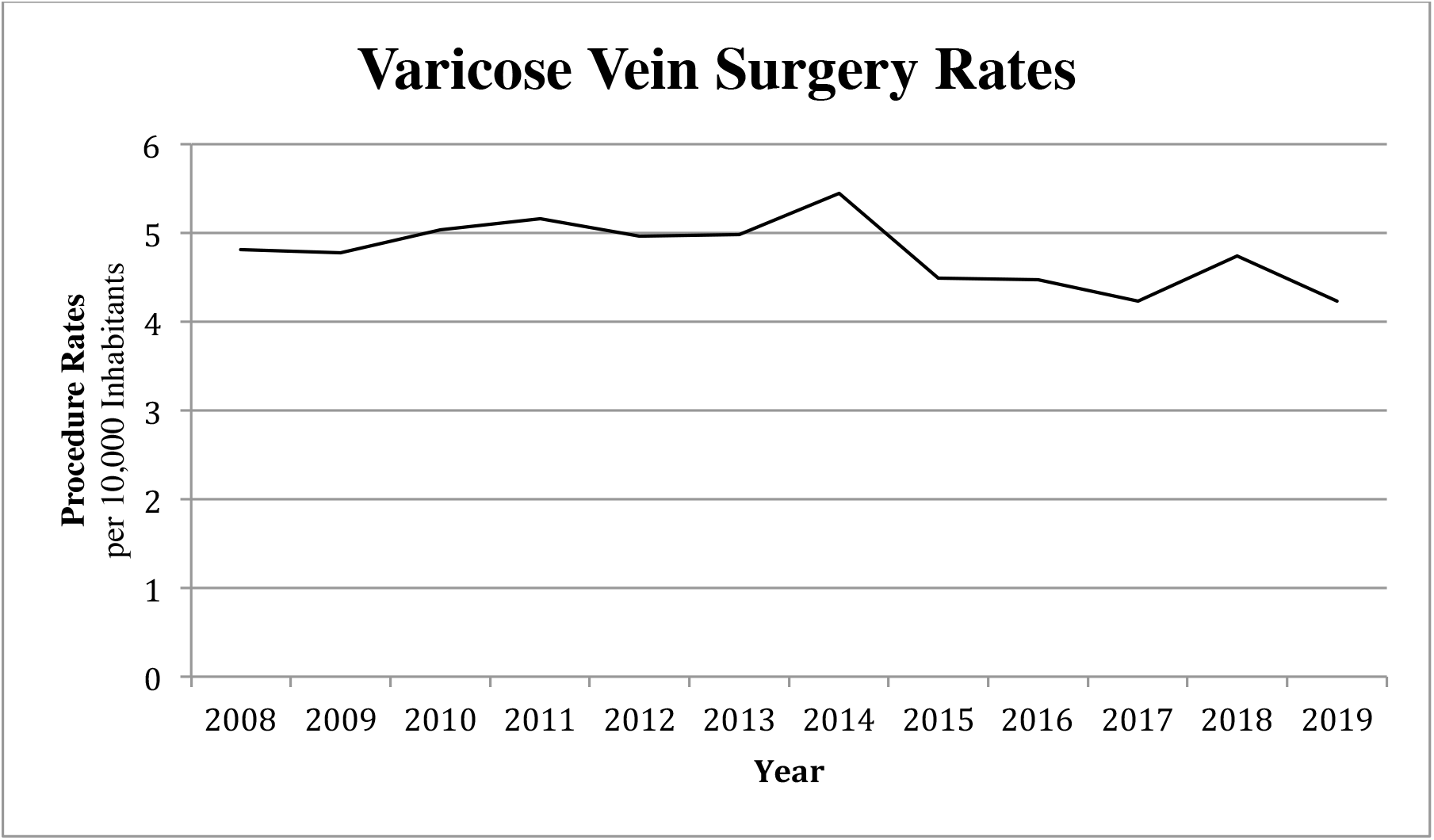
Distribution of procedures rates per 10,000 inhabitants exclusively dependent on the public health system per year.

